# Adaptive Dynamic Social Networks Using an Agent-Based Model to Study the Role of Social Awareness in Infectious Disease Spread

**DOI:** 10.1101/2024.07.16.24310475

**Authors:** Leonardo López, Leonardo Giovanini

## Abstract

The synergy between the spread of infectious diseases and individual behavior is widely recognized. Our pioneering methodology introduces a model based on agents embedded within adaptive temporal networks, providing a nuanced portrayal of daily interactions through an agent-based paradigm. Each agent encapsulates the interactions of individuals, with external stimuli and environmental cues influencing their conduct. Comprising three intertwined elements—individual behavior, social dynamics, and epidemiological factors—the model has been validated against real-world influenza outbreaks, demonstrating superior performance compared to traditional methodologies. Our framework exhibits extensive versatility and applicability by encapsulating individual-level dynamics through elementary rules and simulating complex social behaviors such as social consciousness.

## 1. Introduction

The probability of large-scale disease outbreaks, such as epidemics and pandemics, increases with global connectivity. The Severe Acute Respiratory Syndrome (SARS) outbreak in 2003 (cdc, b), Ebola outbreaks (cdc, a), Asian influenza in 2017 (https://www.facebook.com/WebMD), and *COVID-19* (Kilbourne, 2006) are among the most severe outbreaks in recent history.

Death and illness are the immediate impacts of disease outbreaks, although their socioeconomic effects can be substantial (Barrett et al., 2011a). Depending on the overall state, short-term conditions can induce trends that lead to new social situations. For example, poor economic situations and disease prevalence can lock society into persistent states of poor health and wealth (Bonds et al., 2010). Decease outbreaks and prevalence can also affect government instability. Letendre et al. (2010) analyzed the effects of disease outbreaks on social health and wealth, and their impact on political systems. Persistent disease threats can lead to closing business (Bartik et al., 2020), educational impacts (Rosenthal et al., 2020), disproportionate burden of racial minorities and poor people (Fortuna et al., 2020), general heightened anxiety levels (Vigo et al., 2020) and domestic violence (Kofman and Garfin, 2020).

The essence of epidemic modeling resides in the intricate web of human interactions, influenced by diverse factors including economic conditions, social norms, cultural practices, and individual behaviors. Epidemiologists and healthcare experts employ an array of methodologies to comprehend the complexities of disease transmission dynamics (Barrett et al., 2011b), enabling them to predict and respond effectively to outbreaks. Traditional epidemiological models utilize ordinary differential equations to delineate disease dynamics, categorizing individuals into distinct compartments based on shared characteristics and computing the aggregate movement between these compartments over time (Hethcote, 2000). A plethora of literature offers insights into the application and interpretation of these classical models (Bjørnstad et al., 2020; Cutts et al., 2020; Epstein et al., 2008; Giordano et al., 2020).

Agent-based models (ABM) represent a distinct category of epidemic modeling, where individuals are depicted as agents endowed with unique attributes and behaviors. These agents interact within a simulated environment, generating opportunities for disease transmission. Local interactions give rise to outcomes such as the number of cases, enabling the assessment of outbreak intensity while considering localized characteristics and transmission routes. ABMs offer granularity, allowing the integration of factors like seasonal migrations, spatial distributions, demographics, cultural practices, and individual behaviors (Railsback and Grimm, 2019). Research exploring the application of agent-based models spans various domains Adiga et al. (2018); Chen et al. (2016); Hoertel et al. (2020); Kuylen et al. (2020); Yeom et al. (2014).

The spread of diseases within populations underscores the importance of models representative down to the individual level, a crucial aspect of ABMs. These models define the characteristics of individuals including age, gender, marital status, and household income—as well as their activities, such as starting times, duration, and locations, along with their behaviors. Interactions between agents are simulated using contact networks, each imbued with properties that reflect specific circumstances. Disease dynamics unfold on these networks, evolving in response to changes in agents’ health status, behaviors, and public policies. Subsequently, alterations in the network feedback into agents’ behavior and health. Incorporating this co-evolution is essential for accurately modeling disease spread (Barrett et al., 2008). However, it introduces complexity, as individuals’ schedules may vary based on the health status of those they interact with, as well as their demographics, health conditions, needs, social values, and preferences.

Dignum et al. (2020) proposed an *ABM* based on social simulation tools to analyze the social, economic, and health consequences of policy interventions during *COVID-19* pandemic. The agent’s behavior is determined by a balance of the agent’s needs over a set of possible social contexts, given that the agent’s social value system is consistent with the social context.

Bissett et al. (2021) proposed an integrated multi-dimensional approach to simulating epidemics, addressing not only outbreak control but also related social issues like instability and inequality. It includes a theoretical framework, synthetic population generation, social network construction, and disease evolution simulation. This framework does not contemplate adaptive temporal networks or focuses on individual and social behaviors influenced by environmental cues explicitly. On the other hand, is not able to capture individual-level dynamics and complex social behaviors.

López et al. (2020) addressed the co-evolution problem using an adaptive dynamic networks framework where the disease dynamic results from the aggregation of the agents’ behavior over a social network. It is modeled in a modular fashion such that results from aggregating three blocks: *i*) individual behavior, *ii*) social interactions, and *iii*) disease dynamics. In this way, the behavior of each agent is determined by external stimuli, as well as its perceptions and health state. The framework allows modeling different situations (quarantines, multiple strains and public policies, heterogeneity). The novelty of this framework lies on modeling a complex system through the logic of self-organized systems applied to a social network.

Palomo-Briones et al. (2022) proposed an *ABM* model that includes the cultural orientation of agents to determine their behavior. It models the double causality between individual behaviors, influenced by cultural orientation, and the evolution of a disease. They used the theory of planned behavior and Bayesian inference to model the decisionmaking processes implicit in an agent’s behavior. A set of simulation experiments was developed to demonstrate the role that cultural orientation plays in the management of an epidemic.

Ventura et al. (2022) proposed a model for epidemic spreading in temporal networks of mobile agents that incorporates local behavioral responses. Susceptible agents are allowed to move away from infected agents in their neighborhood. Recently, Gu et al. (2024) used this model to simulates the spread of the disease in the gathered population by combining the susceptible–infected–susceptible epidemic process with human motion patterns, described by moving speed and gathering preference.

Our focus is the development of an *ABM* spatial framework for computational epidemiology using mobile agents to provide a quantitative understanding of the factors. The proposed model is a development of the one introduced by López et al. (2020). It is based on a self-organized logic with a model of the behavior of individuals that elucidates the relationships between individual behavior, activities, and their location with groups and social dynamics. The structure of this work unfolds as follows: In Section 2, we explore the motivation that underlies our model. Section 3 offers a general description of the model, introducing the conceptual framework. The model is designed as an *ABM* where the behavior of each agent is determined by external stimuli and its perceptions of the environment. The model includes four interacting blocks: *i)* individual behavior, *ii)* activities, *iii)* social behavior, and *iv)* health state, providing a comprehensive representation of daily life interactions and their impact on epidemic dynamics. Section 4 explores the implementation details explaining how the model was translated into a computational framework to simulate the Spanish flu epidemic in Geneva in 1918. Subsection 4.4 outlines the estimation of its parameters for a specific case. It includes a thorough examination of the model’s performance against real influenza epidemic data and a validation process. Section 4.5 shows and analyzes the results obtained with the model, demonstrating its efficacy in reproducing real data. Section 5 shows the model’s versatility to model complex social behaviors, such as social awareness and heterogeneous spatial distribution of agents. Finally, Section 6 summarizes the conclusions derived from this work and suggests potential avenues for future research.

## 2. Modeling framework: Scope and Motivations

Complex systems consist of densely interconnected components with constrained nonlinear feedback. These systems have multiple context-dependent relationships with interactions across various scales and domains. They exhibit self-organizing behaviors that evolve, leading to characteristics such as trajectory dependence, nonlinearities, and emergence. Similar conditions can lead to different outcomes, small inputs can cause large effects, and any intervention may result in unintended consequences (Erten et al., 2017; Scarpino and Petri, 2019).

This type of system can be modeled using ***adaptive co-evolutionary networks***, where nodes represent individuals and links represent interactions between them (Gross and Blasius, 2008). Our objective is to develop tools and methods for constructing adaptive co-evolutionary networks to gain a quantitative understanding of the factors that influence behaviors related to epidemic processes. The methodology includes: *i*) modeling and generating individuals, *ii*) constructing social networks, and *iii*) simulating disease transmission. The purpose of this decomposition (Figure 1) is to simplify the system’s description and clarify the connections between individual behavior (social values, preferences, and needs), social practices, and contagion transmission, while employing the most appropriate tools.

**Figure 1:**
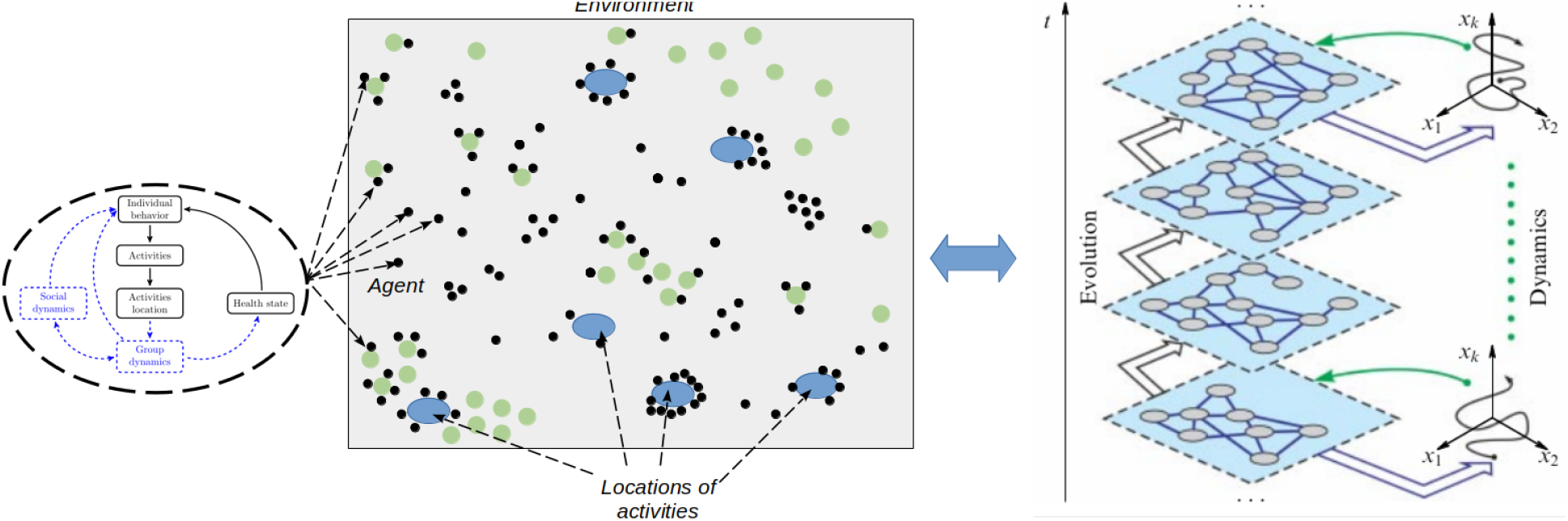
Model diagram and its components

## 3. Modeling framework

There are many challenges in developing *ABM* systems for epidemic modeling. First, we look for a theoretical approach to describe, simulate, and analyze disease dynamics. For this purpose, we use ***graph dynamical systems*** (Mortveit and Reidys, 2007). Second, we seek tools to model populations and the dynamics associated with the disease. They need to represent different features of the behaviors and the activities performed by individuals. They are by nature incomplete and, at different levels of granularity, may be contradictory.

### 3.1 Evolving graph dynamical systems

A graph dynamical system (*GDS*) is an abstract representation of a group of interacting entities (*agents*) and the nature of their interactions (Funke et al., 2019; Mortveit and Reidys, 2007). This representation provides a solid basis to develop models of ***di****ff****usion–reaction processes*** where contagions (viruses, disease, and opinions, among others) are any entity that can propagate through a system.

An ***evolving graph dynamical system*** (*EGDS*) 𝒮 (𝒢, 𝒱, ℰ, 𝒳, *F*_*V*_, *R*) describes a *GDS* that evolve through time. Here *G*(*V, E*_*k*_) ⊆ 𝒢 is the set of graphs with vertex *V* ∈ 𝒱 and edges *E*_*k*_ ∈ ℰ. We use directed dependency graphs, where the direction represent on which vertex the contagion is present. At each sampling time *k* ∈ N_≥0_, each agent *i* ∈ *V* is assigned a state *x*_*i,k*_ ∈ *X* ⊆ 𝒳 ⊆ ℝ^*n*^ is the vertex state set. The system state at time *k* is given by *x*_*k*_ = *x*_1,*k*_, · · ·, *x*_*m,k*_ ^*T*^, where *m* = |*V*| is the number of agents in the system such that the system will have |*V*|^*m n*^ possible states.

Let 𝒩_*k*_[*i*] ∈ 𝒱 be the set of all vertices adjacent to node *i* at time *k*. It is identified from the connectivity graph *G*_*k*_ = *G*(*V, E*_*k*_) by searching of all distance-*r* neighbors. A vertex function *f*_*v*_(*x*_*i,k*_, *x* _*j*∈*N*_*k* [*i*],*k*) ∈ *F*_*V*_ is assigned to each agent *i* that describes its state transition *x*_*i,k*+1_ = *f*_*v*_(*x*_*i,k*_, *x* _*j*∈*N*_*k* [*i*],*k*). The vertex of system agents comprise the sequence *F* = *f*_*v*_(*x*_*i,k*_, *x* _*j*∈*N*_*k* [*i*],*k*) *i V* such that *x*_*k*+1_ = *F*(*x*_*k*_).

The vertex functions *f*_*v*_() ∈ *F*_*V*_ are evaluated in an order determined by *R*. The most common update schemes are ***synchronous*** and ***sequential***. In synchronous update scheme *f*_*v*_() ∀*i* ∈*V* are executed ***simultaneously*** and the system state is given by *x*_*k*+1_ = *F*(*x*_*k*_). In a sequential update scheme, vertex functions are executed ***one-at-a-time*** with the states of all other vertices remaining unchanged. The order in which vertices are updated is determined using one of the possible permutations ? = (*π*_1_, · · ·, *π*_*m*_) ∀*v* ∈ *V* such that the system state is given by *x*_*k*+1_ = *F*_?_(*x*_*k*_), where *F*_?_(·) is the composition of the local functions 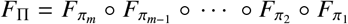 The sequence of states (*x*_0_, *x*_1_,, *x*_*t*_*f*) describes the evolution of the system from initial state *x*_0_ to a specified final time *t* _*f*_ (*x*_*t*_*f*) associated with a sequence of configurations *G*_*k*_ ∀*k* ∈ [1, · · ·, *t* _*f*_].

#### Example

Here we now introduce an example to illustrate the ideas described above. Lopez et al. (López et al., 2020) modeled the transmission of a decease on an evolving contact network built up from a stochastic automaton whose transitions are controlled by a function that determines the transition probability based on the health state, environment, and perceptions of individuals. The changes on the social network are a function of simulation variables such as the duration of contact between infected and susceptible individuals and number of infected, among others. Then, the health component of the vertex function is given by

- If *x*_*v,k*_ = *R* then *x*_*v,k*+1_ = *R* independent of the state of vertex’s neighbors;
- If *x*_*v,k*_ = *I* and 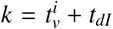 where 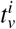 is the time when agent *v* was infected and *t*_*dI*_ is the infection period, then *x*_*v,k*+1_ = *R* otherwise *x*_*v,k*+1_ = *x*_*v,k*_;
- If *x*_*v,k*_ = *S* and ∀ *u* ∈ *N*_*k*_[*v*] that *x*_*u,k*_ = *I* then *x*_*v,k*+1_ = *I* with probability *p*_*v*_ and *x*_*v,k*+1_ = *x*_*v,k*_ with probability (1 − *p*_*v*_) then *x*_*v,k*+1_ = *R* independent of the state of vertex’s neighbors;

The individual-to-individual contact graph is produced by the agent’s position within the cellular automaton, considering a Moore neighborhood of radius *r* ≥ 0, controlled by the individual behavior module. The behavior of agents is determined by factors like perception of the environment, knowledge of the epidemic situation, number of infected individuals, assessments of the epidemiological situation, experiences, feelings, and health state. These factors determine the mobility and dimension of the neighboring zone *r*. The agents move randomly through the cellular automata.

The interaction between health and behavior components of the vertex function determines if the individual would move or not and its relationship with its environment. Then, these actions modify the group of individuals with whom interact, modifying the social network. It is easy to see that the vertex function *f*_*v*_() ∀*v* ∈ *V* is a composition of its components: individual behavior, social behavior, and health state modules.

### 3.2 Framework elements

There has been an increasing interest in understanding how social networks evolve in time. The observation of the network at a specific time represents a state of the system, from which meaningful information can be computed. An ***artificial population*** represents a group of individuals, which can range from a family to the people of a continent. Individuals in an artificial population are endowed with demographic traits (*age, gender, home location*, and *habits*, among others), behaviors and activity patterns where individuals go to particular locations and times.

Typically, these populations are ***not one-to-one*** with targeted populations. Rather, distributions of characteristics of an ariticial population match to those ones of the real one. Thus, the construction of an artificial population is typically one realization of a family of instances. The generation of an artificial population comprises the definition and imputation of the following features:

- **Characteristic of individuals -** Agents have different attributes such as age, gender, income, size, and health conditions;
- **Activities of individuals -** Each agent is assigned an agenda of activities during a day, week, and month, along with their schedule;
- **Locations for activities -** A location is assigned to each activity using all the information and data sources available like commercial, public buildings, leisure, and industrial locations. In case of having no information, a gravity model (locations closer to home) is used;
- **Individual health state -** A disease progression model is assigned to each agent to determine its evolution. It must include information about the disease states, as well as propagation and contagion mechanisms;
- **Individual behavior -** Promotes adaptive responses to problems that individuals face every day (Barrett et al., 2011a). Attitudes and reactions to external stimuli like communication, personal circumstances, and social norms are assigned to each agent; and
- **Social behavior -** Agents interact between them and with the surrounding environment. These interactions are modeled using self-organizing behaviors derived from a Lagrangian framework.

Agents interact with others located in the same places, generating ***groups*** with fixed structures (e.g., agents located at home, workplaces, or regular activities) or time-varying structures (e.g. agents located in transportation hubs, commercial centers, health services, amenity places, or occasional activities). In this context, agents can change their health state, due to the presence of infected agents, modifying their perceptions and behavior, and changing the group’s structure. The resulting vertex functions *f*_*v*_() ∀*v* ∈ *V* is a composition of these three blocks in a given surrounding neighborhood. At a higher level of aggregation, groups interact between them giving rise to ***social dynamics***. Groups behave like agents and aggregate their dynamics at larger time and space scales. The social and group dynamics determine the topology and characteristics of the network, whose nodes are the agents, at the local and global levels respectively. Its evolution depends on the behavior of agents, creating feedback loops between agents (nodes), groups (close networks of other agents), and social (the complete network of agents).

Figure 2 shows Basis Model Unit proposed in this framework. The building blocks and their relationships are denoted in continuous black while the system emergent blocks and their relationships are denoted in dash blue. The social dynamics (time evolution of the network topology) depend on the behavior of agents (dynamic of the nodes), regulated by their health state and group (social) dynamic. In turn, the group (social) dynamic is regulated by the location of activities performed by the agents, which are determined by their behaviors. Then, the social dynamic emerges from the feedback dynamic loops between agents (network nodes) within groups (local network topology) and between groups (global network topology).

**Figure 2:**
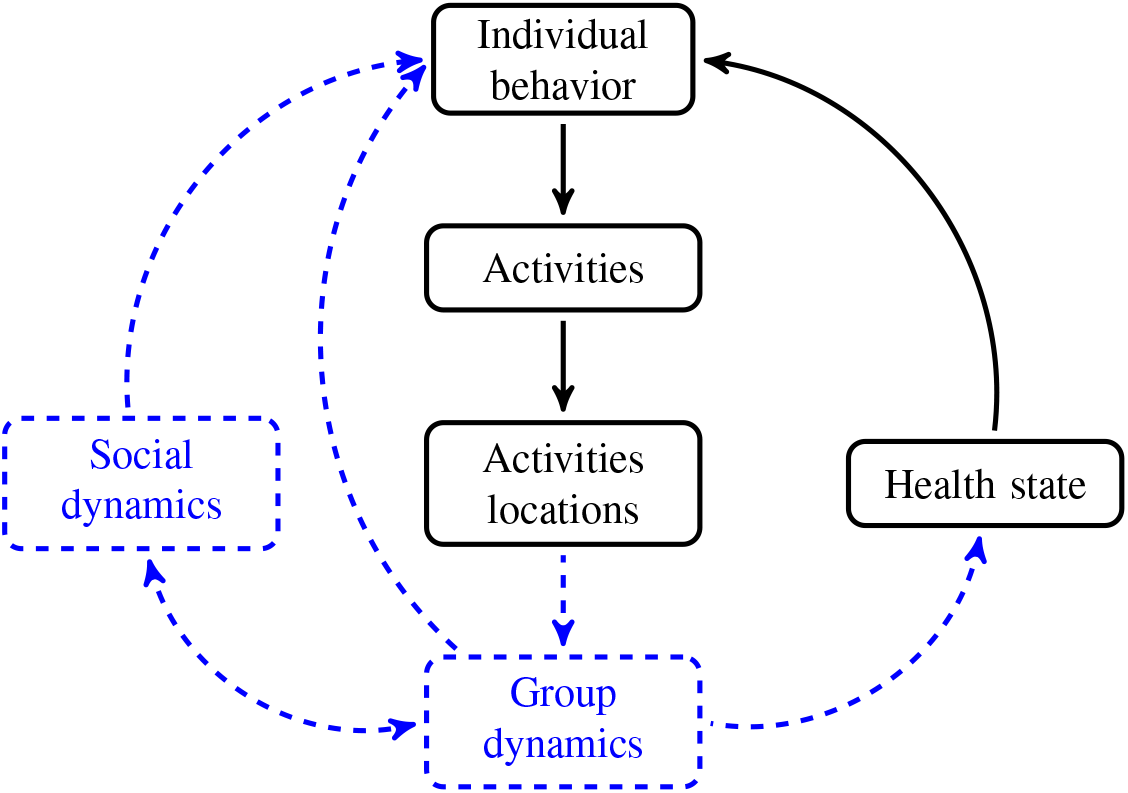
Building blocks of the proposed framework.

Figure 3 shows the connections between the blocks used to model the behavior of agents and the connections with others. The proposed framework links the macro level (***populations***) with intermediate (***group***) and micro level (***agent***) using agents aggregation. The group behavior results from the aggregation of agents in the same location. Using a similar approach, we derive the behavior of a subset of the population by consolidating smaller groups before considering the entire population as a whole.

**Figure 3:**
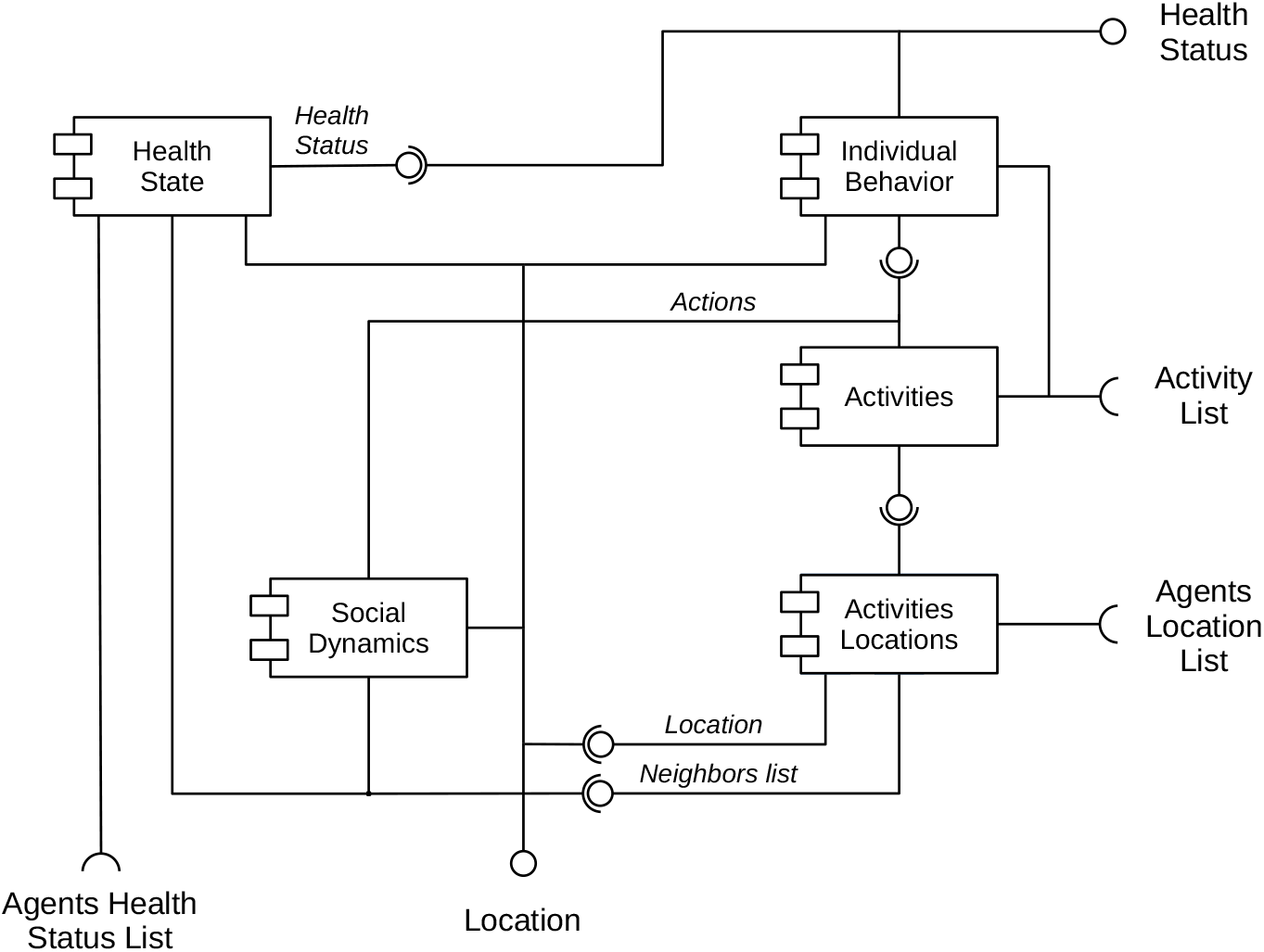
Diagram of the behavior of individuals showing its components and connections

#### 3.2.2. Characteristic of individuals

A baseline population is comprised of different data sources like national census, polls, and surveys produced by the national’s central public statistical services, universities, and international organizations, among others. They provide social, demographic, and economic information about the population and its subdivisions. Census and surveys usually provide data resolved to geographical region levels. For each region, tables of demographic characteristics -like age, gender, and household composition- provide information about their marginal distributions. Then, we must formulate a joint distribution from this information to assemble a population of agents. This distribution is sampled as many times as the number of agents of the model. A similar procedure is followed to generate the households.

If no information about this issue is available, a distribution with *a priory* characteristics is assumed such that satisfies the closest modeling hypothesis to the situation.

#### 3.2.2. Activities of individuals

Data from different sources produced by central national statistical services, universities, and international organizations, among others, are used to set up daily activity sequences for each agent. National census and polls contain detailed information on an agent’s activities throughout a period.

The activity patterns can be modeled with an *extended finite state machine* (*EFSM*). It is an extension of a *finite state machines* that includes additional features like parameters, actions, and guards. They provide better modeling capabilities that allow *EFSM*s to handle complex state-based behaviors (Masek et al., 2018). Parameters support passing information between states, actions enable the execution of specific tasks during state transitions, and guards provide additional conditions for transitions. Within this framework, states encapsulate the range of activities accessible to each individual, inputs denote the variables shaping activity choices, and outputs dictate the specific activity undertaken by the agent at any given moment, thereby dictating the individual’s temporal whereabouts.

Formally, an *EFSM* is defined by the tuple A = (𝒳, 𝒰, 𝒴, *x*_0_, 𝒟, ℒ, ℋ, Υ, Λ), where 𝒳 is a finite state set, *x*_0_∈ is the initial state, is the input set, is the output set, is an *n*-dimensional linear space *D*_1_𝒟 𝒟𝒟7;*D*_*n*_, ℒ is a set of Boolean functions of input variables *l*_*i*_ : *D*→ {0, 1} that defines guard conditions, ℋ is a set of update functions *h*_*i*_ : *D D*, → Υ is transition function Υ : 𝒳 𝒟𝒟7;*D* 𝒟𝒟7;𝒰 → 𝒳 and Λ is the output function Λ : 𝒳 𝒟𝒟7; *D* → 𝒴 The transition function Υ(*x*_*k*_, *u*_*k*_, *l*_*i*_(*D*)) decides what the next state will be *x*_*k*+1_ given the current state *x*_*k*_, input *u*_*k*_ and guards conditions *l*_*i*_(*D*). This means that a transition not only depends on the current state and input but also conditions independent of them. The output function Λ(*x*_*k*_, *h*_*i*_(*D*)) determines which will be the agent’s outputs.

*EFSM*s offer several advantages, compared with other tools for modeling the behaviors of agents:

- **Expressiveness -** It provides a modeling framework that allows modeling complex and flexible state and input-based behaviors;
- **Flexibility -** It can model multiple levels of abstraction and nested behaviors;
- **Complex decision-making -** It can handle a wider range of scenarios and dynamic behaviors, based on adaptable and context-aware conditional transitions and specific tasks execution;
- **Model timing and concurrency -** Actions can be executed after a certain period or transitions can respond to concurrent events.

Activity routines within a household are often interrelated, with the presence of one member influencing the activities of others. For instance, if a child under the age of twelve is at home, there will typically be an adult present. As a result, activity sequences are structured within households to maintain these correlations. Essentially, households are chosen for survey purposes based on their similarity over individual characteristics: a person within a survey is selected that is most similar to a synthetic individual and that survey person’s activities are assigned to the synthetic individual.

#### 3.2.3. Location of activities

Each activity must be located within the modeling space, therefore, methods for designating (*i*) residential addresses, (*ii*) public transport stations, (*iii*) workplaces, (*iv*) shopping centers, and (*v*) recreational facilities must be implemented.

Home locations can be assigned using data produced by national and regional statistical services, governmental agencies, and private organizations. Similarly to previous Subsections, home locations can be selected by similarity of individual characteristics between data and synthetic individuals, measured with distribution and set distance measures. Web mapping platforms contain information about residential locations, streets, road maps, and public transport networks.

Locations of workplaces, commercial centers, amenities, and businesses for individuals can be assigned combining information from data produced by governmental and private organizations, as well as web mapping platforms. If no information about their locations, they can be assigned using gravity models or uniformly. The concept involves assigning locations for activities based on a probability that is directly relative to the capacities of the buildings and inversely related to the distances from the current location..

#### 3.2.4. Individual health state

The health status of an agent dictates its epidemiological condition throughout the epidemic process. This aspect captures the disease progression within agents (***intra-host***). Various tools can be employed to model this progression, depending on the specific modeling requirements and the type of information available and exchanged. Differential equations, automaton, probabilistic models, and immune-response models are among the possible tools.

One effective tool for modeling health state dynamics is the *EFSM*, which offers a robust approach for capturing the intricate state-based behavior of disease progression. *EFSM* enables the combination of timed deterministic and probabilistic transitions. It supports multiple co-circulating diseases, various strains, disease co-factors, and sophisticated interventions, such as contact tracing and antiviral reserves. Moreover, *EFSM* facilitates the modeling of various epidemiological scenarios, including isolation measures, immunization efforts, and multiple strains, by adjusting the states and transitions accordingly, all while maintaining computational simplicity.

#### 3.2.5. Individual behavior

Human behavior is affected by countless factors including interpersonal communication, emotions, personal beliefs, and perceptions. Responses towards diseases are determined by a combination of these factors, choosing among the available alternatives. Different models have been proposed to represent this component of the behavior of individuals (Anderson et al., 2004; Carruthers, 2011; Dao-Ping et al., 2007).

Emotions are mechanisms that allow individuals to respond to stimuli from the environment. They overpower the decision-making process to select a suitable reaction when there is too much information to process or too little time to react. Emotions evolved to maintain the functional balance of individuals by counteracting the information and energy flow by reducing their effect on them. Emotions like pain, anger, nervousness, security, relaxation, and excitement, can be seen as self-regulatory homeostatic mechanisms (Kowalczuk and Czubenko, 2016). They are the result of a complex chain of connected events with stimuli, including feelings, psychological changes, impulses to action, and goal-directed behaviors (Plutchik, 2001).

There are two main theories to describe the creation (or triggering) of emotions. ***Appraisal theory*** gives preponderance to cognitive processes over emotions, such that emotions emerge from the analysis of stimuli through cognitive processes (Lazarus, 1991). On the other hand, ***somatic theory*** gives preeminence to emotions over cognitive processes, such that an individual can immediately invoke emotions associated with specific events without analyzing the stimuli. Surveys on modeling emotions can be found in (Hieida and Nagai, 2022; Van Haeringen et al., 2023), among others.

Emotional states can characterized in terms of their similarity, intensity, and duration, among other parameters.

Emotions can be classified into three levels (Xiao-Juan and Wei-Ren, 2007):

- **Primary emotions -** are individuals’ intrinsic responses to external stimuli. They are connected with both somatic (spontaneous physical feelings, dependent on specific stimulus) and appraisal (associated with objects, places, or situations from previous experiences) theories. They last from a few seconds to minutes;
- **Secondary emotions -** emerge when primary emotions are linked to both current and past perceptions and experiences. These emotions can be consciously recognized and verbalized. They are associated with both theories, potentially intertwined with motivational factors. These emotions exhibit slow changes and can persist from a few minutes to several weeks; and
- **Senior emotions -** are those produced by the course of long-term social contacts in a given environment (Damasio, 1994). They are based on personality and last years.

In literature, one can find many works concerning the issue of modeling human emotions: *CBI* (Marsella, 2003), *EMA* (Gratch and Marsella, 2004), *FLAME* (El-Nasr et al., 2000), *FearNot!* (Dias et al., 2014), *Thespian* (Si et al., 2006), *Peactidm* (Marinier III et al., 2009) and *Wasabi* (Becker-Asano and Wachsmuth, 2010), among others. However, none of these models have being used in the epidemiological modeling context.

Fuzzy Cognitive Maps (*FCM*) provide a powerful tool for modeling systems in terms of interacting concepts, that represent a state or a characteristic, and linkages, that express their relationship, in a hierarchical structure (Karyotis et al., 2018). A *FCM* incorporates both the experience and the accumulated knowledge of the system, which is derived from the expertise of individuals who understand the system’s operation and its components under various conditions, often based on statistical analysis. In this model, concepts are represented as nodes, and the connections between these nodes depict the causal relationships among them. These maps model a collection of concepts and the cause-effect relationships between them. Interconnections between different concepts *C*_*i*_ and *C*_*j*_ are characterized by a weight *w*_*i j*_, which describes the degree of causality and influence between them (Mei et al., 2014). Weights can take values within interval [−1, 1] such that their values quantify the causality and their sign indicates the type: **positive** when *w*_*i j*_ *>* 0 (*C*_*i*_ increases then *C*_*j*_ increase) and **negative** when *w*_*i j*_ *<* 0 (*C*_*i*_ decreases then *C*_*j*_ decreases).

#### 3.2.6. Social behavior

In multi-agents systems, groups of agents interacts and evolve within the environment. Their simulation allows direct evaluation of agents’ behaviors and interactions (as well as groups and systems) in response to changes in the agents and the environment (Epstein, 2006). Properties at agent’s level correspond to the characteristics, behaviors, health status, and activities of the agent. Group-level (local) properties match to the characteristics, behaviors, health state, and activities of a group of agents defined by their neighborhood. They emerge from the aggregation of the agents’ properties as a result of the interaction between them and the environment. Finally, system-level properties are the global properties of the environment in which agents live and emerge as a result of the interaction between groups (Epstein, 2006).

In his seminal papers, Reynolds (Reynolds, 1987, 1999, 2000) proposed a consensus-based algorithm that has been widely studied for modeling multi-agent systems. In this context, agents make use of information exchange between them to reach a common value (known as a ***consensus***) for cohesive behavior as a group. Following this idea, multi-agent systems can be modeled as a group of *m* agents with a simple dynamic

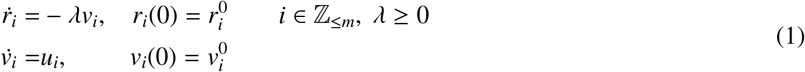

where *r*_*i*_ = (*x*_*i*_, *y*_*i*_)^*T*^ is their position in space, 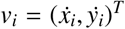 their velocity and 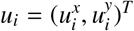 is their input that modifies their behavior (Olfati-Saber, 2006). The input *u*_*i*_ is given by

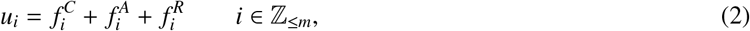

where 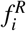 is a term that model the local behavior of agent *i* through a short-range random movement, 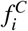 term that quantifies the interactions of agent *i* with agents in its neighborhood

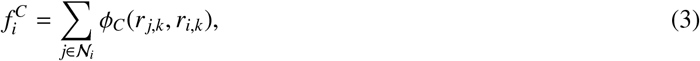

where 𝒩_*i*_ denotes the group of neighbors and *ϕ*_*C*_(*r* _*j,k*_, *r*_*i,k*_) is the consensus term with the agents in the neighboring zone (see Figure 4). The term 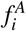 quantifies the interactions of agent *i* with the associated locations of activities and environment

**Figure 4:**
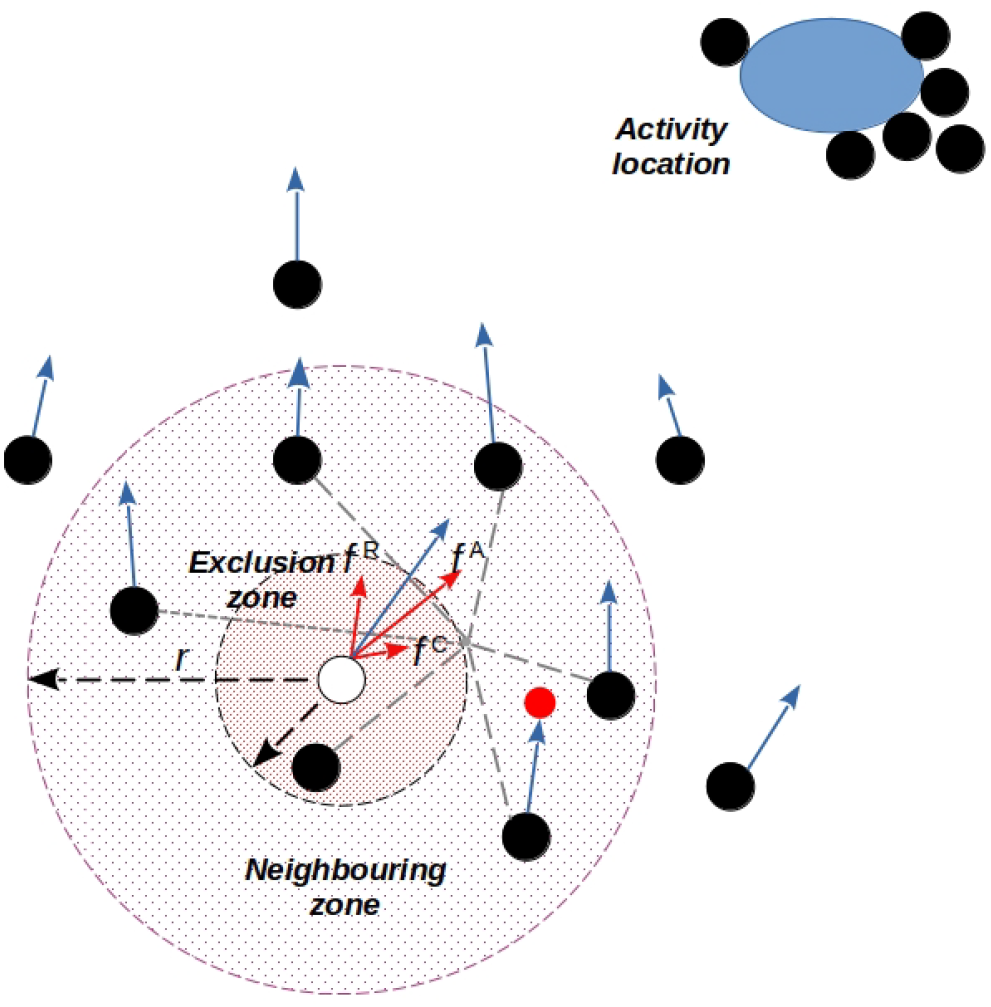
Illustration of agents’ behavioral components.

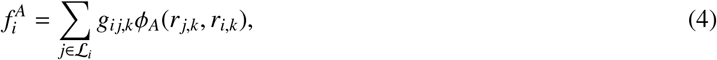

where ℒ_*i*_ denotes the set of activities locations for agent *i, ϕ*_*C*_(*r* _*j,k*_, *r*_*i,k*_) is the consensus term with activities locations, represented as rendezvous points, and *g*_*i j,k*_ is an output of the Individual behavior block that indicates which activity is performed or not (*g*_*i j,k*_ = 1 or *g*_*i j,k*_ = 0). Agents representing locations and obstacles are configured with predetermined behaviors. For instance, stationary agents depict obstacles and fixed locations (Figure 4).

Social behavior is determined by the day-to-day routines of individuals, which is broken down into smaller time scales like hours and divided into social activities and non-social activities like staying at home. This behavior is modeled through an *EFSM* whose states represent the activity that the agent is going through. In this sense, the *EFSM* serves as an intermediate layer between the individual behavior and the group dynamics (and the social dynamics), affecting how individuals move through the grid (Figure 1). Individual behavior takes into account the influence of other agents and the environment; and their objectives through the function *g*_*i j,k*_.

Then, the behavior of the systems arises from the aggregation of the behaviors of the agents that compose it (selforganization). The idea of self-organization rests on the premise of ***individual rationality*** (Cristiani et al., 2015; Lee, 2011), a doctrine in which each agent pursues the best possible outcome for himself. In this way, each agent tries to do the best for himself taking into account the effects of the actions of the rest on itself. Then, the behavior of the system results from the superposition of the behaviors of agents, the emergent behavior of the system, which is more complex than the simple aggregation of the behaviors of agents (Helbing et al., 2011).

The collective behavior of individuals within groups arises from their individual actions and interactions with others. These interactions are modeled using feedback functions, which are governed by finite state automata with probabilistic transitions. These transitions, determined by the time variable, dictate the movement or actions of individuals. Further elaboration on this topic will be provided in subsequent sections.

## 4. Epidemiological simulation

The modeling framework proposed in Section 3 is used to model the Spanish flu epidemic in the Swiss canton of Geneva in 1918 (Chowell et al., 2006) and explore the effects of agents behavior on the spread of influenza over the population. The interaction network emerges from the aggregation of the behavioral building blocks (individual behavior, activities, and health state) of agents interacting between them and with the surrounding environment. The agents and locations serve as nodes within the evolving structure of the social network, where interaction patterns fluctuate in response to shifts in an agent’s health condition (e.g., opting to stay home when unwell) and behavior (e.g., minimizing non-essential activities during an outbreak), alongside public interventions (e.g., school closures, vaccination drives). Consequently, this dynamic network influences individual health outcomes, such as minimizing exposure to infectious agents outside the home or increasing contact with infected individuals within it.

The processes of between-host transmission and within-host progression are intricately linked yet computed independently. Between-host transmission initiates the within-host disease progression, transitioning the agent’s health state from susceptible to infected. The progression of the disease is entirely dictated by the health state (a component of the vertex function) governing within-host progression. It is modeled with a stochastic *EFSM* that supports multiple co-circulating diseases and heterogeneity, among other features. On the other hand, disease transmission is determined by the health state of agents within a neighborhood (group dynamic), governed by the aggregation of agents’ behavior and locations of activities performed by them (the other two components of the vertex function). The agent behavior is modeled using a *FCM*, which is able of representing their behavior toward the main external stimuli of the environment. The sequence of activities is modeled using a stochastic *EFSM* such that we model agents’ habits (deterministic transitions) with unexpected events. All of this is also affected by the demographics of individuals (e.g. agent’s income and susceptibility to disease influence their decision on work, and physical needs and confidence influence their decision to practice sports).

### 4.1 Individual health state

Agent’s epidemic state is defined as: *EFSM 𝒜* = (𝒳, *𝒰, 𝒴, x*_0_, *ϒ*, Λ) where 𝒳 is a finite set of states, *u*_*i*_ ∈ 𝒰 ⊆ ℝ _[0,1]_ *i* ∈ *𝒩*_*v*_ is the probability of contagion between agents *v* and *I* ∈ 𝒩_*v*_ and *y*_*v,k*_ ∈ 𝒴 ⊆ ℝ _[0,1]_ is the infection rate such that *y*_*v,k*_ = *u*_*i,k*_ ∈ 𝒩_*v*_. The set of epidemic states, denoted as 𝒳, encompasses six distinct conditions: *S* for susceptible, *E* for exposed, *I* for infectious symptomatic, *A* for infectious asymptomatic, *R* for recovered, and *D* for deceased or unoccupied. The initial state vector *x*_0_ of each agent is assigned in a probabilistic way such that the number of infected (symptomatic and asymptomatic), exposed, and susceptible is equal to the initial conditions of the epidemic. The infection process transitions from *E* to *I* or *I*, developing after an incubation period *τ*_*I*_. On the other side, the recovery process occur after the recovery period *τ*_*R*_. The output *y*_*v,k*_ = Λ(*x*_*v,k*_) = *ωx*_*v,k*_ computes the infection rate only if the agent is in state *I* (*ω* = *β*) or *A* (*ω* = *q*), otherwise *ω* = 0.

The diagram presented in Figure 5 illustrates the state transition graph and parameters defining the epidemic state within the *EFSM*. Here, *β* represents the transmission probability, *ρ* denotes the proportion of reported infectious individuals, *γ* signifies the recovery rate, *α* indicates the diagnosis rate, *q* represents the infection rate among asymptomatic individuals, *N*_*e*_ represents the initial population of exposed individuals, *N*_*i*_ signifies the initial population of infectious individuals, while *τ*_*I*_ and *τ*_*R*_ correspond to the incubation and recovery periods, respectively.

**Figure 5:**
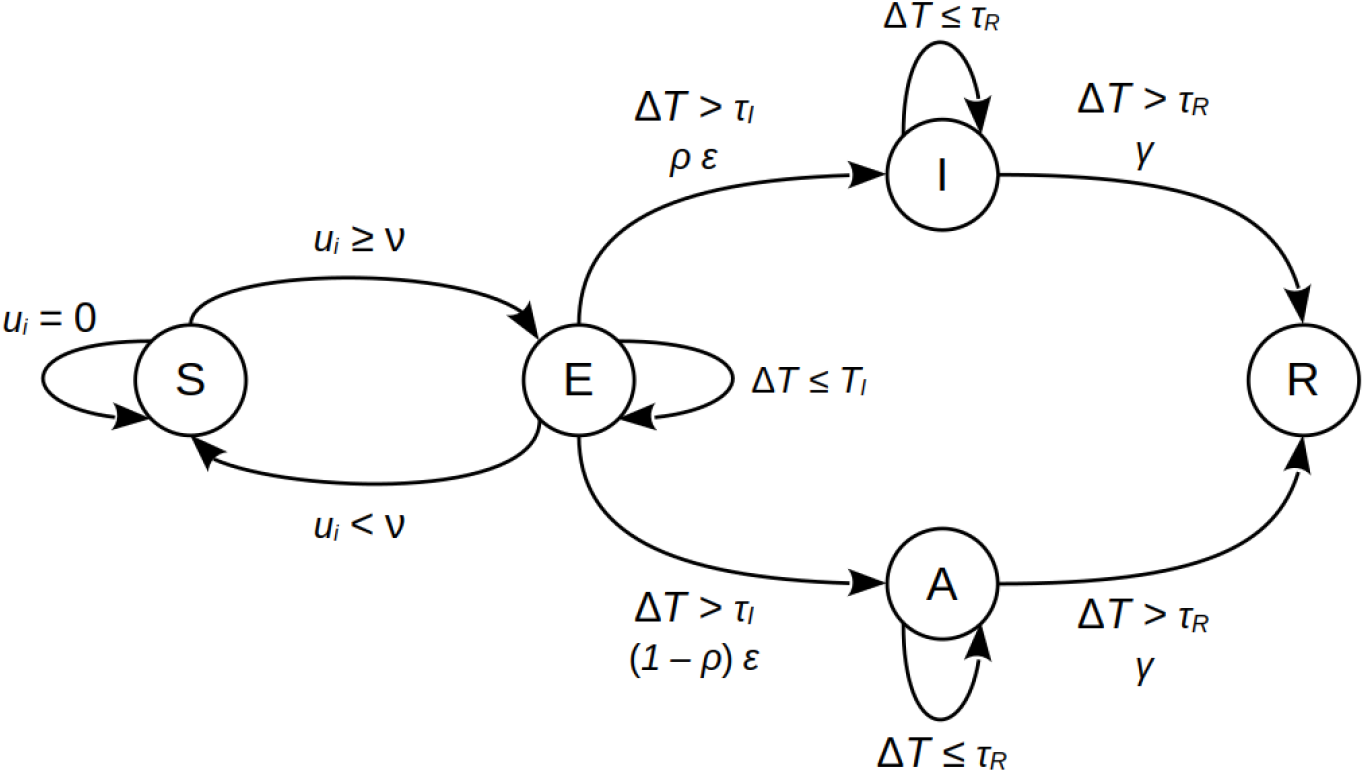
State graph of the epidemic states model

When an agent is in state *S*, it receives an active input *u*_*i*_ ∈ 𝒰if agent *i* ∈𝒩_*v*_ is in either state *I* or *A*. This input signifies the probability of between-host transmission resulting from interactions between agents *v* and *i*. The state transition function, denoted as ϒ: 𝒳 → 𝒴, is applied to *x*_*v,k*_, probabilistically determining *x*_*v,k*+1_. This function operates in two stages: one governing state changes due to infection and recovery, and another governing movement. State changes are determined using two probability matrices: one for transitions under no input (Table 1), where *µ* represents the probability of natural death and *γ* represents the probability of recovery, and another for transitions resulting from contact with infectious individuals (Table 2).

**Table 1:** Probabilistic transition matrix derived from classical model parameters (López et al., 2020), reflecting an aggregation of individual probabilities in large populations. The model explores variations in spatial distributions and population sizes.

| | $S$ | $E$ | $I$ | $A$ | $R$ |
| --- | --- | --- | --- | --- | --- |
| $S$ | $1 - \mu$ | 0 | 0 | 0 | 0 |
| $E$ | 0 | $1 - (\varepsilon + \mu)$ | 0 | 0 | 0 |
| $I$ | 0 | 0 | $1 - (\gamma + \mu)$ | 0 | $\gamma$ |
| $A$ | 0 | 0 | 0 | $1 - (\gamma + \mu)$ | $\gamma$ |
| $R$ | 0 | 0 | 0 | 0 | $1 - \mu$ |
| $D$ | $\mu$ | $\mu$ | $\mu$ | $\mu$ | $\mu$ |

**Table 2:** Matrix representing transitions between agent states upon contact occurrence.

|  |  |  |
| --- | --- | --- |
| | $S$ | $E$ |
| $S$ | $1 - \lambda/\nu$ | $\lambda/\nu$ |
| $E$ | $0$ | $1$ |

The derivation of the transition matrix originates from classical model parameters (Chowell et al., 2006). While deterministic in nature within the classical model, they manifest as an aggregation of individual probabilities, predicated on the assumption of a sufficiently large population. Consequently, it is the probabilistic transition that underpins the classical model, rather than vice versa. Direct application of ϒ to each agent reveals a diminishing variability with population growth, ultimately converging with the deterministic evolution of the classical model. Moreover, the initial distribution of individuals within the environment may be random, conforming to the hypothesis of homogeneous distribution in large population sizes. Additionally, the model offers exploration into various spatial distributions and population sizes.

The output function Λ : 𝒳 → 𝒴 ∈ 𝒟 computes the infection rate based on an individual’s state: *I*(*ρ, β*) or *A*(1 *ρ, β*), where *ρ* ∈ *ℝ*_[0,1]_ denotes the probability of infection upon contact with an asymptomatic individual. This probability, distinct from the classic model’s *β*^′^, represents the likelihood of transmission through contact. The potential number of infectious contacts *c* is influenced by the neighborhood size, governing connectivity with other network nodes, albeit not directly correlating with the contact count. The infection rate for each infectious individual arises from the interplay between the *β*^′^ probability and the potential number of contacts *c*. The *β* value, derived from parameter settings, is determined by the neighborhood size selection and the ^*β*ss′^ parameter from the classical model, computed as 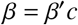.

### 4.2 Social behavior

Social behavior plays a pivotal role in shaping individual interactions, exerting a significant influence on the dynamics and structure of social networks. Utilizing self-organization principles rooted in Lagrangian concepts (Mogilner et al., 2003; Cucker and Smale, 2005; Olfati-Saber, 2006), the social behavior of agents is modeled. Here, individual behaviors define the collective behavior of the system, illustrating emergent properties arising from the aggregation of individual actions. Consequently, an agent’s actions are not solely influenced by local cues but also by the collective information within the system. One approach to defining behavior is through the rules proposed by Olfati-Saber (Olfati-saber, 2006).

- Avoid collision with neighboring agents;
- Minimize interactions with neighboring agents; and
- Move to target locations.

To obtain these behaviors, the consensus terms *ϕ*_*C i*_(·) and *ϕ*_*A i*_(·) of input *u*_*i*_ (equation 2) are given by

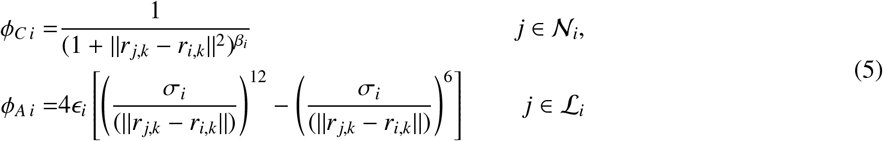

where *β*_*i*_ = 0.5, *σ*_*i*_ = 1.0 and *ϵ*_*i*_ = 1.0. The consensus terms associated with activities *ϕ*_*A j*_(·) *j* ∈ *ℒ*_*i*_ are modulated by the output of the *EFSM* that models the activities of the agents through *g*_*i j,k*_.

Since we have no information about the activities performed by individuals at Geneva in 1918, we will assume a simple activity model: agents can move to the nearest social center (*SC*) (i.e. schools, offices, public transport stations, among others) and then come back to home (*H*), or remain in the same place (*R*), home or social center. In other words, they determine where they move to.

Figure 6 summarizes the *EFSM* that models the activities performed by agents. For simplicity, it was decided to model the activities of agents at social centers (*SC*) or their homes (*H*). When agents move from a state of rest (*M*), they stay in the same place but move randomly in the vicinity (due to *f* ^*R*^ component of social behavior). The output of the *FCM* affects the decision of performing (or not) a given activity, determining if agents move toward a place or not (cancel or no contact with other agents). In this way, the activities performed by agents are determined by time-dependent transition functions that determine the following states (activity) in the *EFSM*. These time constraints are modeled through the guard, timing, and concurrency conditions of the states.

**Figure 6:**
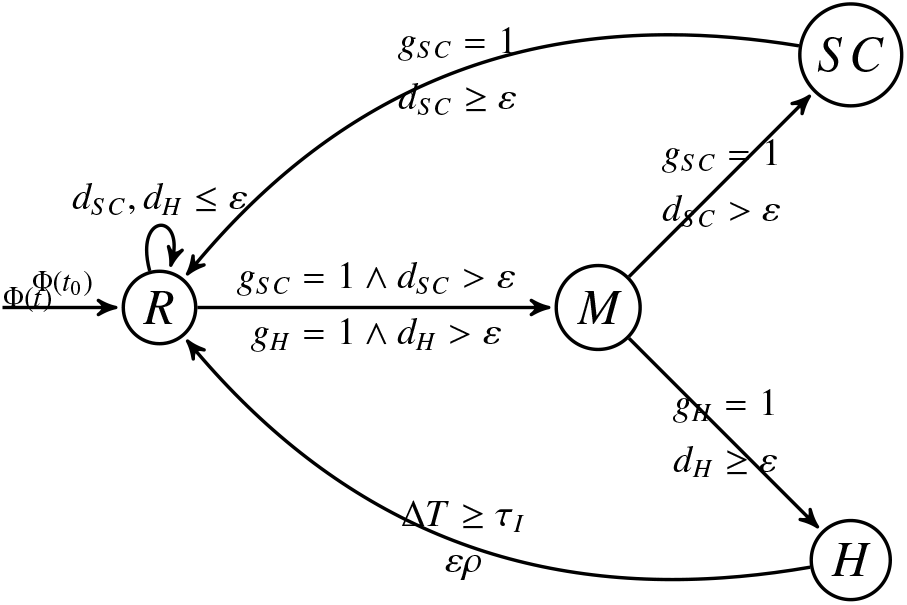
Model for agents activities.

### 4.3 Individual behavior

Individual behavior is captured using a *FCM* framework, following the model introduced by Mei et al. (2014). In this model, concepts *C*_*i*_ *i* = 1, · · ·, 10 are categorized into three distinct groups:

#### Input concepts

serve as model inputs, where *C*_1_ denotes proximity to infected individuals, *C*_2_ represents proximity to recovered individuals, and *C*_5_ signifies awareness of the global epidemic situation, reflecting the environmental cues perceived by the agent (primary emotions).

#### Internal concepts

encapsulate the individual’s emotions and feelings, encompassing secondary emotions. Here, *C*_3_ denotes the individual’s health state (as determined by epidemic behavior), *C*_4_ signifies awareness of the local epidemiological situation, *C*_6_ represents the assessment of both local and global epidemiological conditions, *C*_7_ indicates the level of optimism, *C*_8_ denotes memory of similar situations, and *C*_9_ encompasses instant reactions.

#### Output concept

corresponds to senior emotions (*C*_10_), representing the actions undertaken by the agent following a decision-making process.

The inputs of the model are *u*_1_ the density of infected agents that have contact with the agent, *u*_2_ the density of recovered individuals that have contact with the agent, *u*_3_ the health state of the agent, and *u*_4_ the knowledge of the local epidemiological situation. The value of *C*_10_ quantifies the individual’s perception of the overall epidemiological situation, controlling the size of the neighboring zone (*C*_10_ = *r*) and *C*_11_ control the transitions of the activities block of the agent.

Figure 7 illustrates the connections between the concepts *C*_*i*_ and inputs *u*_*i*_, along with their corresponding weights. The *C*_10_ value constrains the number of contacts an individual can make within the environment, thus influencing the control entry *u*_*i*_(*t*). At each iteration, the values of input concepts *C*_1_ and *C*_2_ are estimated based on the number of infected and recovered individuals in the agent’s vicinity. The value of concept *C*_3_ is determined by entry *u*_3_ and reflects the agent’s health state transition.

**Figure 7:**
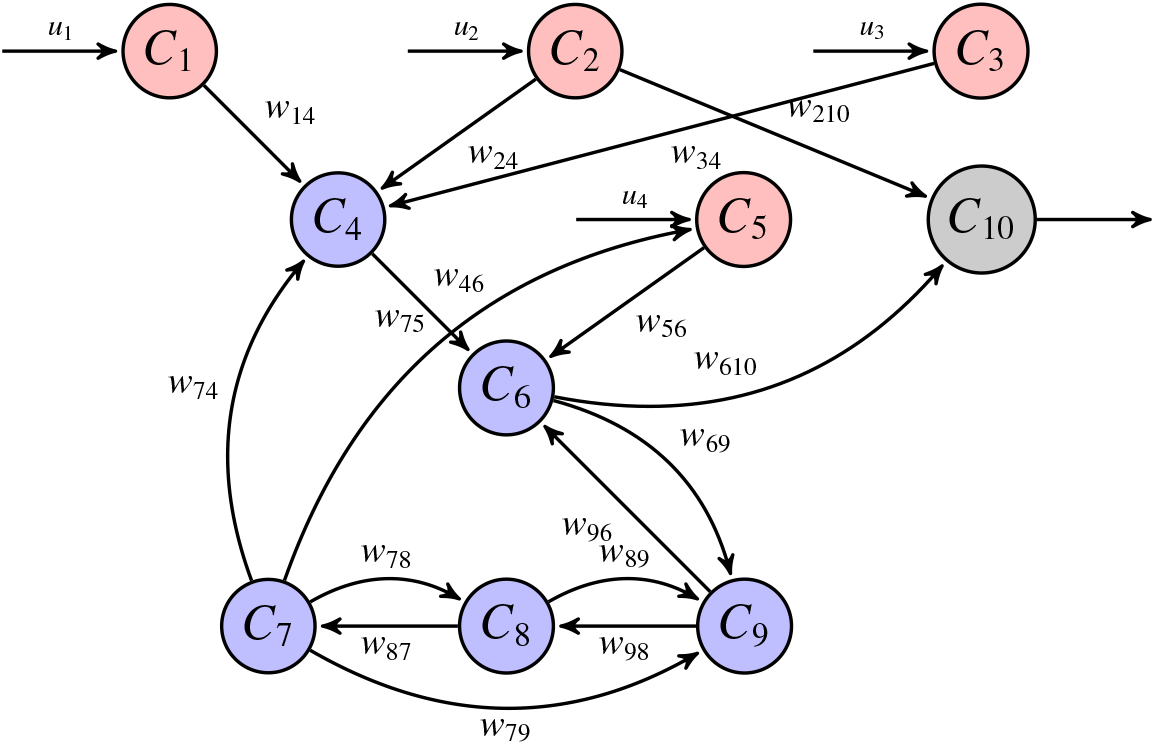
Diagram of the *FCM*.

#### Model fitting

To assess the model’s efficacy, we examine the impact of the Spanish flu outbreak in the Swiss canton of Geneva in 1918 (Chowell et al., 2006). Our parameter estimation process involves a meticulous two-step approach. Initially, a comprehensive ***global search*** across the parameter space is conducted to identify promising candidate regions, employing stochastic optimization techniques such as simulated annealing (Deb et al., 2002). Subsequently, a ***local search*** is performed within each candidate region using gradient-based optimization algorithms (Byrd et al., 2000) to pinpoint the optimal parameters. Leveraging stochastic optimization methods ensures robust initial points for gradient-based optimization techniques. The objective function employed in this process is the normalized square error.

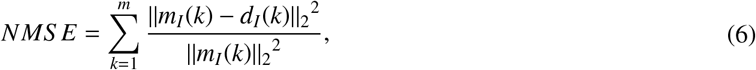

The variable *m*_*I*_(*k*) represents the predicted number of infected agents (i.e., infected individuals) by the model, while *d*_*I*_(*k*) corresponds to the epidemiological data collected during the outbreak. Table 3 presents the estimated parameters of the proposed model.

**Table 3:** Model parameters.

| $\beta$ | $\rho$ | $\gamma$ | $\alpha$ | $q$ | $\tau_I$ | $\tau_R$ | $N_e$ | $N_i$ |
| --- | --- | --- | --- | --- | --- | --- | --- | --- |
| 8.1 | 0.085 | 0.246 | 0.465 | 0.3 | 2.3 | 6.8 | 207 | 136 |

To calibrate the *FCM* algorithm proposed by Mei (Mei et al., 2014), we utilized training data. The computation of each concept *C*_*i*_ takes into consideration the influence of other concepts *C*_*j*_ *j* ≠ *i*, along with the causal relationships between them. Given that our model focuses on a localized outbreak (at the city level), assigning *C*_5_ = 0.5 equates to a phase 4 alert status as per the World Health Organization guidelines. This phase is characterized by confirmed instances of human-to-human transmission within the community.

In the training of the *FCM*, another crucial aspect to consider is the choice of threshold function utilized to update concepts *C*_*i*_. While the sigmoid function is a common choice for this purpose, it’s important to note that its domain and range are bounded. To ensure consistency across the inference interval, a linear function is employed, preserving the same slope throughout. This approach is particularly advantageous for modeling individual behavior during the Spanish flu outbreak. Por supuesto, aquí tienes la notacioón en LaTeX para la ecuacioón revisada:

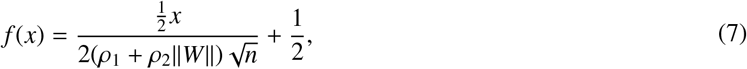

where *ρ*_1_ and *ρ*_2_ represent values within the range of [0, 1], *n* denotes the total number of concepts, and *W* denotes the weight matrix defined as

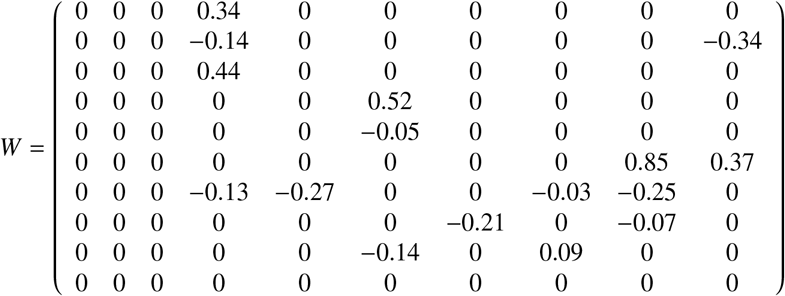

### 4.5. Model validation

Figure 8 shows the simulation results provided by the *SEIR* model developed by Chowell et al. (Chowell et al., 2006), the *ABM* model proposed by Lopez et al. (López and Rodó, 2020) and the one proposed in this work. The average behaviors of both *ABM* models are estimated from 1000 numerical simulations. Figure 9 shows the residuals of the models along the epidemic process (Figure 9.a) and their distributions (Figure 9.b), allowing to evaluate their ability to reproduce the epidemic dynamic.

**Figure 8:**
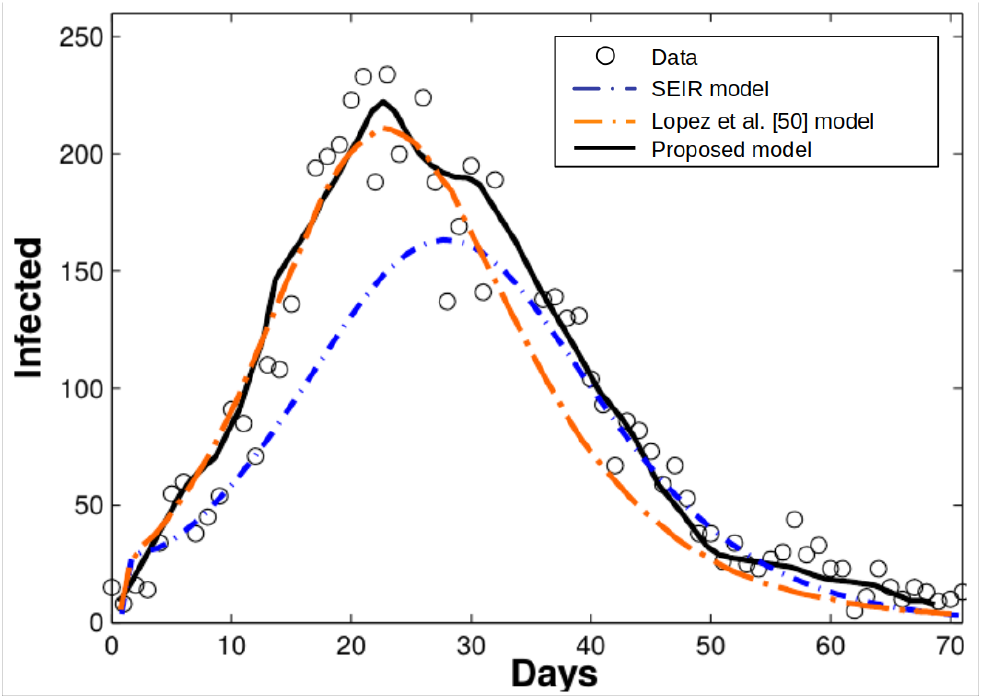
Average responses of the models and real data.

**Figure 9:**
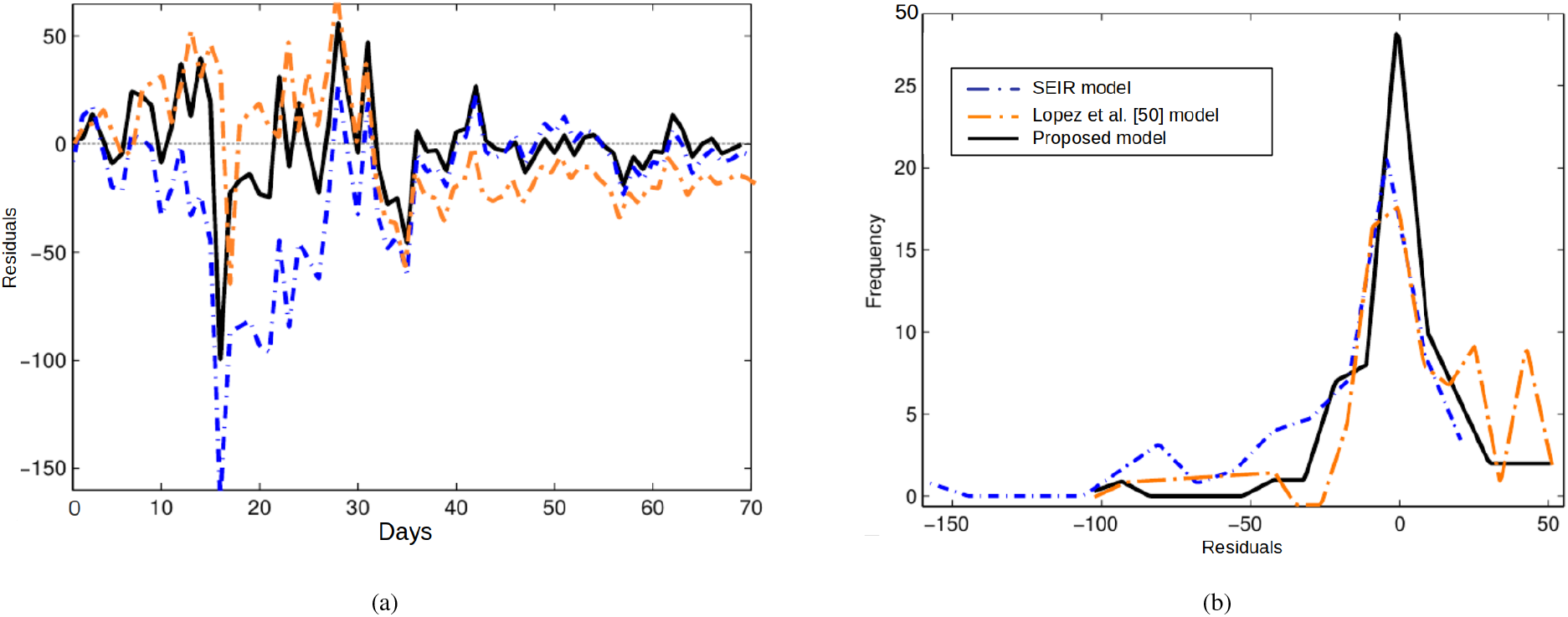
(a) Residuals of the model and (b) distribution of residuals.

Figure 8 shows the results for each model. Only the proposed model captures the epidemic dynamic along the entire process. The number of infected agents grows rapidly until reaches the peak (day 23), then it decreases until vanished with two different velocities: In the period after the peak (days 30 to 50) the number of infected falls from 228 to 22, and then it falls from 22 to 10 (days 50 to 70) in similar periods.

The *ABM* model proposed by López et al. (2020) captures the initial phase up to the peak (days 0 to 30), estimating correctly the time and magnitude of the peak. However, it fails to reproduce the epidemic data in the final phase (days 30 to 70). Finally, the *SEIR* model only captures the end phase of the epidemic process (days 35 to 70). It fails to reproduce the epidemic behavior in its initial phase (days 0 to 35) and to estimate the time and magnitude of the peak (see Figure 9.a).

Figure 9.a shows the residual distribution of overtime. The larger residuals occur around the peak (15 ≤*k* ≤35 days), while the initial (0≤ *k* ≤ 15 days) and final (35 ≤*k* ≤70 days) phases have smaller magnitudes. During this period, the data show higher variability due to the irregularity of reporting cases. The magnitude of the residuals generated by the proposed model is lower than the ones generated by the other models. Only 10% of its samples are larger than ±25 and 3% larger than ±50. The *ABM* model of Lopez et al. (López and Rodó, 2020) generates 20% of residuals larger than ±25 and 6% larger than ±50. Finally, the *SEIR* model generates 25% of residuals larger than ±25 and 10% larger than ±50.

Figure 9.b shows the residual distribution of each model. Their statistical parameters are shown in Table 4, where 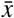 denotes the mean, *M*_*d*_ the mode, *std* the standard deviation and *S* the bias coefficient computed using the second Pearson coefficient formula.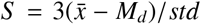 The distribution of the proposed model has a similar mean 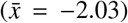 and mode (*M*_*d*_ = −1.83) with a nil bias coefficient (*S* = −0.06). The skewness coefficient shows a symmetric distribution of the residuals (*skew* = −0.12) although it is peaky (*kurt* = 2.12). On the other hand, the residuals distribution of the *SEIR* and *ABM* model of Lopez et al. (López and Rodó, 2020) are biased and offsetting in different directions: the *SEIR* residuals are biased toward the left, while the *ABM* of Lopez et al. towards the right of the distributions mean.

**Table 4:**
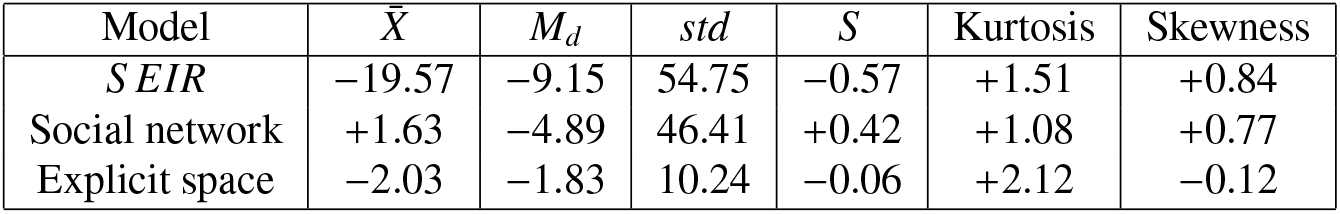
Statistics of residuals.

The model was also numerically validated using the *Akaike information criterion*

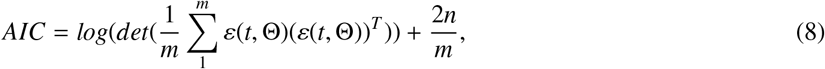

where Θ is the set of *n* uncertainty parameters, *m* is the number of simulations or samples, and *ε*(*t*, Θ) is the error measure. It measures the model quality considering accuracy and complexity simultaneously. It is widely used to measure the quality of models and validate them (Symonds and Moussalli, 2011), being equivalent to a cross *k-leaveone-out* method in longitudinal data models validation (Fang, 2011). The resulting coefficients are *AIC* = 7.5 for the *SEIR* model, *AIC* = 6.1 for the networked *ABM* model, and *AIC* = 3.8 for the proposed model.

The residuals of the models were computed using *NMS E* function defined in (6), resulting in a *NMS E* = 3.3 for the *SEIR* model, a *NMS E* = 1.6 for the *ABM* model of Lopez et al. (López et al., 2020) and a *NMS E* = 1.05 for the proposed model. These results were computed from the data and the average behavior of 1000 simulations for each model.

Finally, the statistical significance of these results was assessed by comparing the probability of residuals. For this test, statistical independence of error adjustment for different data sets is assumed and the errors of binomial distributions are approximated with a Gaussian distribution. Three new datasets for each model were generated with gaps (data points removed) in the data used to estimate the parameters chosen with uniform probability: *i* one dataset with 10 data points removed, *ii* the other with 20 data points removed and *iii* the last one with 30 data points removed. The parameters of each model were estimated from each data set following the same procedure like the described in Subsection 4.4. Then, 1000 simulations were executed for each parameter set to gather a reliable approximation of the average response and the corresponding residuals. Finally, we tested the hypothesis *P*(*Error*_*model*_ *< Error*_*S EIR*_) *> p* was tested for each parameter set. Table 5 presents the results.s

**Table 5:**
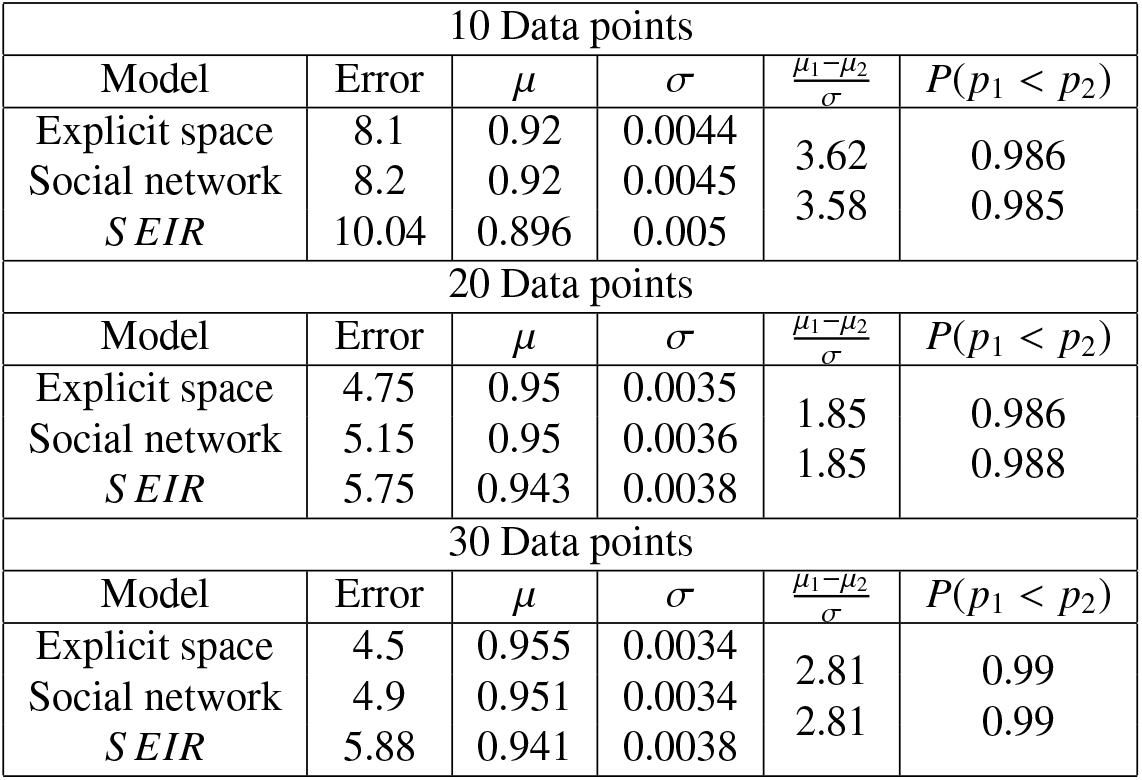
Errors significance for models.

| 10 Data points |  |  |  |  |  |
| --- | --- | --- | --- | --- | --- |
| Model | Error | $\mu$ | $\sigma$ | $\frac{\mu_1 - \mu_2}{\sigma}$ | $P(p_1 < p_2)$ |
| Explicit space | 8.1 | 0.92 | 0.0044 | 3.62 | 0.986 |
| Social network | 8.2 | 0.92 | 0.0045 | 3.58 | 0.985 |
| SEIR | 10.04 | 0.896 | 0.005 |  |  |
| 20 Data points |  |  |  |  |  |
| Model | Error | $\mu$ | $\sigma$ | $\frac{\mu_1 - \mu_2}{\sigma}$ | $P(p_1 < p_2)$ |
| Explicit space | 4.75 | 0.95 | 0.0035 | 1.85 | 0.986 |
| Social network | 5.15 | 0.95 | 0.0036 | 1.85 | 0.988 |
| SEIR | 5.75 | 0.943 | 0.0038 |  |  |
| 30 Data points |  |  |  |  |  |
| Model | Error | $\mu$ | $\sigma$ | $\frac{\mu_1 - \mu_2}{\sigma}$ | $P(p_1 < p_2)$ |
| Explicit space | 4.5 | 0.955 | 0.0034 | 2.81 | 0.99 |
| Social network | 4.9 | 0.951 | 0.0034 | 2.81 | 0.99 |
| SEIR | 5.88 | 0.941 | 0.0038 |  |  |

## 5. Results and discussion

This section assesses the capabilities of the proposed framework to model different scenarios by analyzing the effects of the agent’s epidemiological awareness and initial population distribution on the epidemic dynamic.

### 5.1. Perception of epidemic situation

Interactions are essential for comprehending epidemic development, severity, and progression. The proposed model offers valuable insights into the underlying mechanisms driving epidemic spread by shedding light on the nuanced dynamics resulting from the interplay between perceptions and actions. To illustrate how the individual behavior block shapes the epidemic dynamics, the agents’ perception of an epidemic is modified through *C*_5_, a composite measure of the information registered from media channels like television, radio, newspapers, and social platforms.

The values of *C*_5_ for each agent were obtained from a normal distribution for each simulation. Firstly, agents **indi**ff**erence** to epidemic were modeled using a *C*_5_ mean value of 0.15. Then, agents **concern** about epidemic were modeled using a *C*_5_ mean value of 0.85. Finally, agents showing a balance between caution and disregard, like in a **normal scenario**, were considered through a *C*_5_ mean value of 0.5. Figures 10 show the average behaviors of 500 simulations for each scenario, where agents were uniformly distributed at the beginning of the simulation.

**Figure 10:**
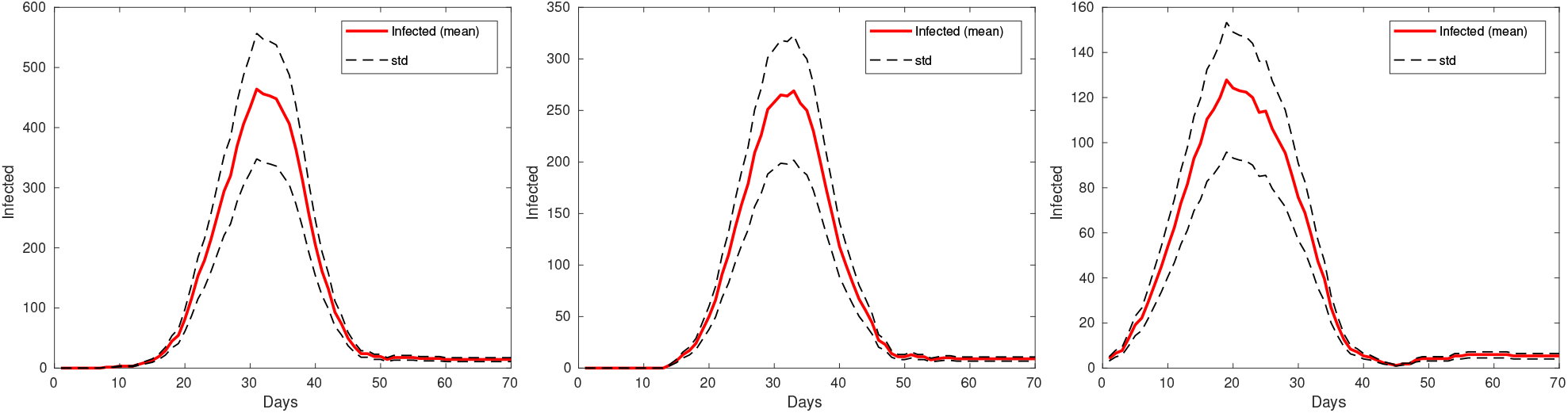
Average response and standard deviation for (*a*) indifferent, (*b*) normal and (*c*) concerned agents’ attitude.

Figure 10.a shows the behaviors for an indifference scenario (*C*_5_ = 0.15). The peak is significantly higher than the real response (see Figure 8) with a delayed start and shorter epidemic process. This behavior is consistent with the assumptions since agents show high mobility, amplifying the likelihood of effective contact and the daily tally of infections. Figure 10.b shows the collective behavior for a normal scenario. Remarkably, it closely mirrors the dynamic behavior depicted in Figure 8, particularly in the similarity of the peak of cases. This observation emphasizes the pivotal role of individual perception in shaping overall epidemic dynamics (Abdulkareem et al., 2020). Finally, Figure 10.c shows the average collective behavior for a cautious scenario (*C*_5_ = 0.85). As it was anticipated, the peak is significantly lower than the real response (see Figure 8) with a similar duration and a small secondary wave after day 45. These behaviors are consistent with the assumptions since individuals reduce their mobility, decreasing the probability of effective contact.

### 5.2 Distribution

By relaxing the assumption of homogeneity, we uncover behaviors that deviate significantly from the ideal scenario. Specifically, when initially infected agents occupy a small portion of the interaction region a noticeable decrease in the epidemic incidence is observed: the peak happens earlier and its magnitude is lower. However, the epidemic process lasts longer, and a secondary wave appears. This phenomenon can be attributed to the heterogeneity of agent distribution that slows down the transmission dynamics.

Figures 11 show the average behaviors of 500 simulations with different initial distributions of infected agents. The number of initial infected agents is 0.1% of the population. One set of simulations where performed without the individual behavior block (without *FSM*), such that they move according to daily activities. In this configuration simulations show a similar number of infected agents at the peak (around 400 agents), the process begins earlier and ends in similar periods (around day 50).

**Figure 11:**
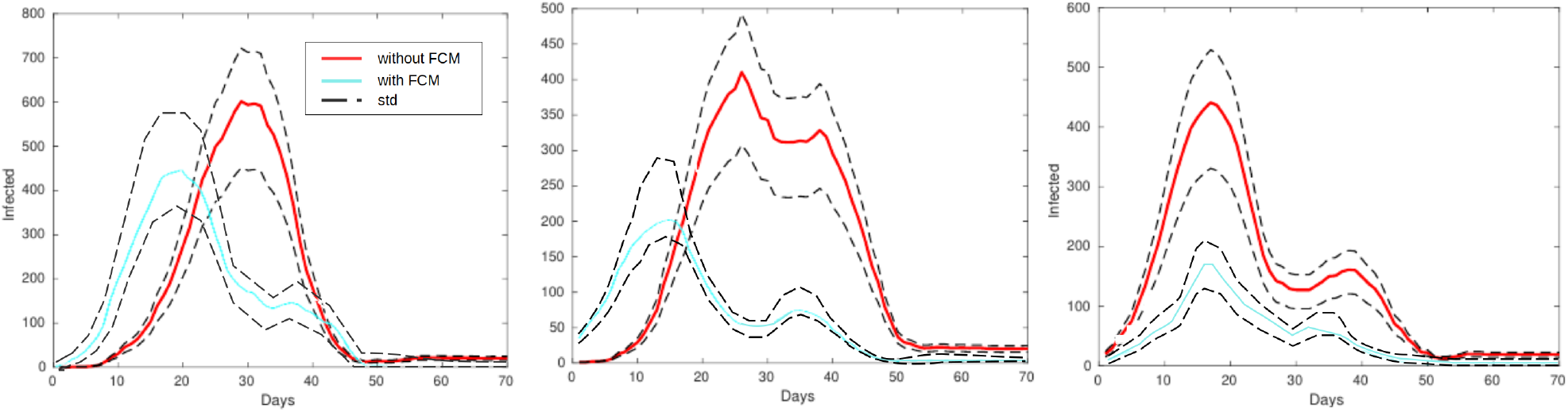
Average response and standard deviation for (*a*) full, (*b*) a 1*/*4 and (*c*) 1*/*8 distribution of initial infected agents.

The other set of simulations where performed including the effect of perceptions on the behavior of the agents (with *FSM*), such that they move combining daily activities with perception. In this configuration simulations show a lower number of infected agents, the process begins earlier and ends in similar periods (around day 50). Once again, we noted the impact of delayed recovery, attributed to individuals’ tendencies to distance themselves from groups, thereby opting for voluntary isolation. This behavior underscores the significance of individuals’ awareness of the epidemic context in their immediate surroundings, leading to a heightened sense of paranoia and social avoidance phenomenon facilitated by the integration of *FCM* into individual behaviors.

In essence, our model is able of capturing the complex dynamics of individual interactions, ranging from cohesive group formations to voluntary isolation, under varying degrees of epidemic awareness. These findings underscore the model’s robustness in simulating realistic scenarios of epidemic spread, providing valuable insights into the complex interplay between individual behaviors and epidemic dynamics.

## 6. Conclusions

Our study introduces an innovative approach for modeling epidemic dynamics by employing a contact network framework that not only captures temporal dynamics but also elucidates the social dimension of outbreaks through interactions among network actors. Our framework offers a transparent and straightforward implementation, demonstrating its capability to accommodate and to replicate real-world situations with remarkable precision. This multiscale modeling approach facilitates the fitting of epidemic dynamics, akin to classic population-based models. Once fitted, epidemiological parameters can be uniformly assigned to individuals, enhancing model consistency across different contexts.

The integration of behavioral aspects via the individual behavior block enhances the model’s ability to capture the dispersion of data across epidemic waves, particularly toward the epidemic’s conclusion. Our model accurately replicates epidemic outbreaks, effectively reproducing the rapid surge in infected individuals at the epidemic’s onset. Notably, it adapts well to varying input distributions for global epidemic perception, as demonstrated in Figures 10 and 11 showcasing its robust performance under different scenarios.

The use of a Lagrangian formulation of self-organization to model social behavior significantly enhances effective contact between susceptible and infectious individuals, especially in homogeneous distributed environments. Additionally, the output of *FCM* significantly influences individuals’ interactions, capturing both rapid dynamics at the epidemic’s onset and the emergence of smaller outbreaks as conditions improve.

Understanding and incorporating individual behavior and activities are crucial in epidemic modeling, as human behavior significantly influences disease dynamics. Our comprehensive approach, considering individual behaviors, emotions, and social interactions, contributes to a better understanding of disease spread and aids in the development of effective public health interventions. In summary, our study advances epidemiological research by providing a versatile modeling framework that improves epidemic predictions and enhances our understanding of the impact of individual behaviors on disease transmission dynamics.

Furthermore, to enhance the model’s realism and reflect real-life scenarios more accurately, future work should focus on incorporating mechanisms of individual learning. In reality, individuals learn from previous experiences, which significantly influences their behavior and decision-making processes. To address this aspect, various options can be explored, such as language models or deep learning techniques, among others. By integrating learning mechanisms, the model can better capture the adaptive nature of human behavior in response to evolving epidemic situations. Additionally, the methodology presented in this study warrants validation with other diseases where behavioral factors play a crucial role, such as Tuberculosis. This validation process will provide further theoretical grounding and ensure the applicability and robustness of the model across different epidemiological contexts.

## Data Availability

All data produced in the present study are available upon reasonable request to the authors

## Acknowledgments

The authors wish to express their gratitude to the National University of Litoral (UNL) and the Research Institute for Signals, Systems, and Computational Intelligence (sinc(*i*)), Santa Fe, Argentina, as well as the National Scientific and Technical Research Council, Argentina, for their support. This work has been funded by the National Scientific and Technical Research Council.

## Authors statement

**Leonardo López**: Conceptualization, Data curation, Formal analysis, Investigation, Methodology, Validation, Visualization, Writing – original draft. **Leonardo Giovanini**: Conceptualization, Formal analysis, Funding acquisition, Investigation, Methodology, Supervision, Validation, Writing – original draft

